# Association between early discontinuation of endocrine therapy and recurrence of breast cancer among premenopausal women in a Danish population-based cohort

**DOI:** 10.1101/2020.10.13.20212217

**Authors:** Lindsay J Collin, Deirdre P Cronin-Fenton, Thomas P Ahern, Michael Goodman, Lauren E McCullough, Lance A Waller, Anders Kjærsgaard, Per Damkier, Peer M Christiansen, Bent Ejlertsen, Maj-Britt Jensen, Henrik Toft Sørensen, Timothy L Lash

**Affiliations:** Department of Epidemiology, Rollins School of Public Health, Emory University, Atlanta, GA, USA; Department of Clinical Epidemiology, Aarhus University Hospital, Aarhus, Denmark; Department of Surgery, The Robert Larner, M.D. College of Medicine at The University of Vermont, Burlington, VT, USA; Department of Biostatistics and Bioinformatics, Rollins School of Public Health, Emory University, Atlanta GA, USA; Institute of Pathology, Aarhus University Hospital, Aarhus, Denmark; Department of Clinical Biochemistry and Pharmacology, Odense University Hospital, Odense, Denmark; Department of Clinical Research, University of Southern Denmark, Odense, Denmark; Department of Plastic and Breast Surgery, Aarhus University Hospital, Aarhus, Denmark; Danish Breast Cancer Group, Rigshospitalet, Copenhagen University Hospital, Copenhagen, Denmark; Rigshospitalet, Copenhagen, Denmark

## Abstract

**Purpose:** Premenopausal women diagnosed with estrogen receptor (ER) positive breast cancer are prescribed 5–10 years of endocrine therapy to prevent or delay recurrence. Many women who initiate endocrine therapy fail to complete the recommended course of treatment. In this study, we evaluated the association between early discontinuation of adjuvant endocrine therapy and breast cancer recurrence in a cohort of premenopausal women.

**Patients and Methods:** We identified 4,503 premenopausal ER+ breast cancer patients who initiated adjuvant endocrine therapy and were registered in the Danish Breast Cancer Group clinical database (2002–2011). Women were excluded if they had a recurrence or were lost to follow-up less than 1.5 years after breast cancer surgery. Endocrine therapy was considered complete if the patient received at least 4.5 years of treatment or discontinued medication less than 6 months before recurrence. Exposure status was updated annually and modeled as a time-dependent variable. We accounted for baseline and time-varying confounders *via* time-varying weights, which we calculated from multivariable logistic regression models and included in regression models to estimate hazard ratios (HR) and accompanying 95% confidence intervals (CI) associating early discontinuation with breast cancer recurrence.

**Results:** Over the course of follow-up, 1,001 (22%) women discontinued endocrine therapy. We observed 202 (20%) recurrences among those who discontinued endocrine therapy, and 388 (11%) among those who completed the recommended treatment. The multivariable-adjusted estimated rate of recurrence was higher in women who discontinued endocrine therapy relative to those who completed their treatment (HR=1.67, 95% CI 1.25, 2.14).

**Conclusion:** These results highlight the importance of clinical follow-up and behavioral interventions that support persistence of adjuvant endocrine therapy to prevent breast cancer recurrence.

## Background

More than two-thirds of premenopausal women diagnosed with breast cancer have estrogen receptor (ER)-positive disease and are therefore candidates for endocrine therapy.^1^ Current guidelines recommend that premenopausal women diagnosed with ER-positive tumors receive adjuvant tamoxifen therapy for at least five years post-diagnosis to prevent or delay a recurrence.^2^ Tamoxifen is a selective ER modulator with metabolites that compete with estrogen at the ER binding site.^3^ Tamoxifen was first introduced in the adjuvant setting in 1989 with treatment guidelines recommending one year of adjuvant therapy.^4^ As evidence from clinical trials accumulated, treatment guidelines have extended the intended duration of adjuvant tamoxifen therapy from one year, to two years, to five years.^5^ A pooled analysis of tamoxifen clinical trials among breast cancer patients diagnosed with ER-positive disease reported risk ratios (RR) for the effect of one, two, and five years of tamoxifen therapy versus placebo of 0.79, 0.72, and 0.50, respectively.^6^ Since 2013, a duration of up to ten years of tamoxifen is recommended for premenopausal women diagnosed with non-metastatic breast cancer.^7^ These changes in guidelines reflect accumulating evidence of the benefit of longer duration tamoxifen therapy.

Barriers to tamoxifen treatment adherence and completion include sociodemographic characteristics suggestive of poor access to resources, concurrent medication use, disease severity, and overall health status of the patient.^8–13^ Lower treatment completion has been observed in younger patients (aged less than 40 years), unmarried patients, and those with a low level of social support.^14^ Switching hormonal therapies can also reduce adherence.^15,16^ Endocrine therapies sometimes cause adverse side effects (*e.g*. hot flashes, nausea, fatigue, arthralgia, vaginal bleeding, and fractures) that can affect a patient’s choice to continue therapy.^17,18^

Early discontinuation and poor adherence to prescribed therapies may contribute to poor patient outcomes. In the US, approximately 50% of breast cancer patients never complete the intended duration of endocrine therapy.^19^ In Denmark, the proportion of patients completing adjuvant therapy is higher. A population-based study using the Danish Breast Cancer Group (DBCG) clinical database reported that approximately 84% of postmenopausal breast cancer patients in Denmark completed endocrine therapy, which was defined as medication use for at least 4.5 years.^20^ No study has quantified the effect of early discontinuation of adjuvant endocrine therapy on breast cancer recurrence in an exclusively premenopausal cohort. In this study, we quantify the association between early discontinuation of endocrine therapy and breast cancer recurrence among premenopausal women with a stage I–III first primary breast cancer diagnosis in Denmark.

## Methods

### Study Population

Denmark provides universal tax-funded health care with residency-based entitlement and availability of government-maintained nationwide registries, providing longitudinal sources of routinely collected administrative, health, and clinical quality data.^21^ The Predictors of Breast Cancer Recurrence (ProBe CaRe) cohort is a study of premenopausal women diagnosed with breast cancer in Denmark and reported to the DBCG.^22^ The cohort includes 4,600 women diagnosed with stage I–III ER-positive primary breast cancer between January 1, 2002 and December 31, 2011. Women were excluded if they had a previous cancer diagnosis. As breast cancer recurrences that occur within the first 1.5 years following breast cancer surgery are unlikely to be related to endocrine therapy, we also excluded 97 women in the cohort (68 of whom were diagnosed with a breast cancer recurrence) who had less than 1.5 years of follow-up. The final study population included 4,503 premenopausal women diagnosed with ER-positive breast cancer. Members of the cohort had follow-up information available until August 31, 2019, with recurrence follow-up from 1.5 years after surgery until the first of a) recurrence, b) death, c) 10 years of follow-up, d) loss to follow-up due to emigration, e) another incident malignancy, or f) the end of available follow-up. Mortality and emigration were identified using the Danish Civil Registration System, which is updated daily.^23^ Emigration or withdrawal from clinical follow-up were the only sources of loss of follow-up, which have impacted less than 5% of the study population. This study was approved by the Danish Data Protection Agency (record number KEA-2017-4), the Danish Medicines Agency, the DBCG (DBCG-2014-15), and the local Institutional Review Boards of the US co-investigators. This study is based on routinely collected registry data, so Danish law does not require an ethical board’s approval in Denmark, nor does it require patient consent to participate.

### Exposure: Early Discontinuation

According to the DBCG guidelines, women diagnosed with breast cancer are recommended to undergo semi-annual examinations during the first 5 years after diagnosis and annual examinations in years 6–10.^24^ Women undergoing treatment for breast cancer receive endocrine therapy at the treating hospital at each follow-up visit. Treatments prescribed during these follow-up visits are registered by the DBCG, which documents whether a treatment was received and the type of therapy (tamoxifen or aromatase inhibitors). At each visit, a 6-month supply of endocrine therapy is dispensed.

Early discontinuation of endocrine therapy was defined as receipt of endocrine therapy for less than 4.5 years. If a recurrence, emigration, or death occurred before 5 years elapsed after treatment initiation, then discontinuation of therapy was deemed early if therapy was discontinued more than half a year before the event. Initiation of endocrine therapy was defined as the first date of receipt of endocrine therapy according to the DBCG registry. Discontinuation of endocrine therapy was based on the last date of receipt of endocrine therapy, with consideration of the 6-month supply of endocrine therapy prescribed. **Figure 1** illustrates the cohort entry, induction period, and follow-up considerations.

**Figure 1.**
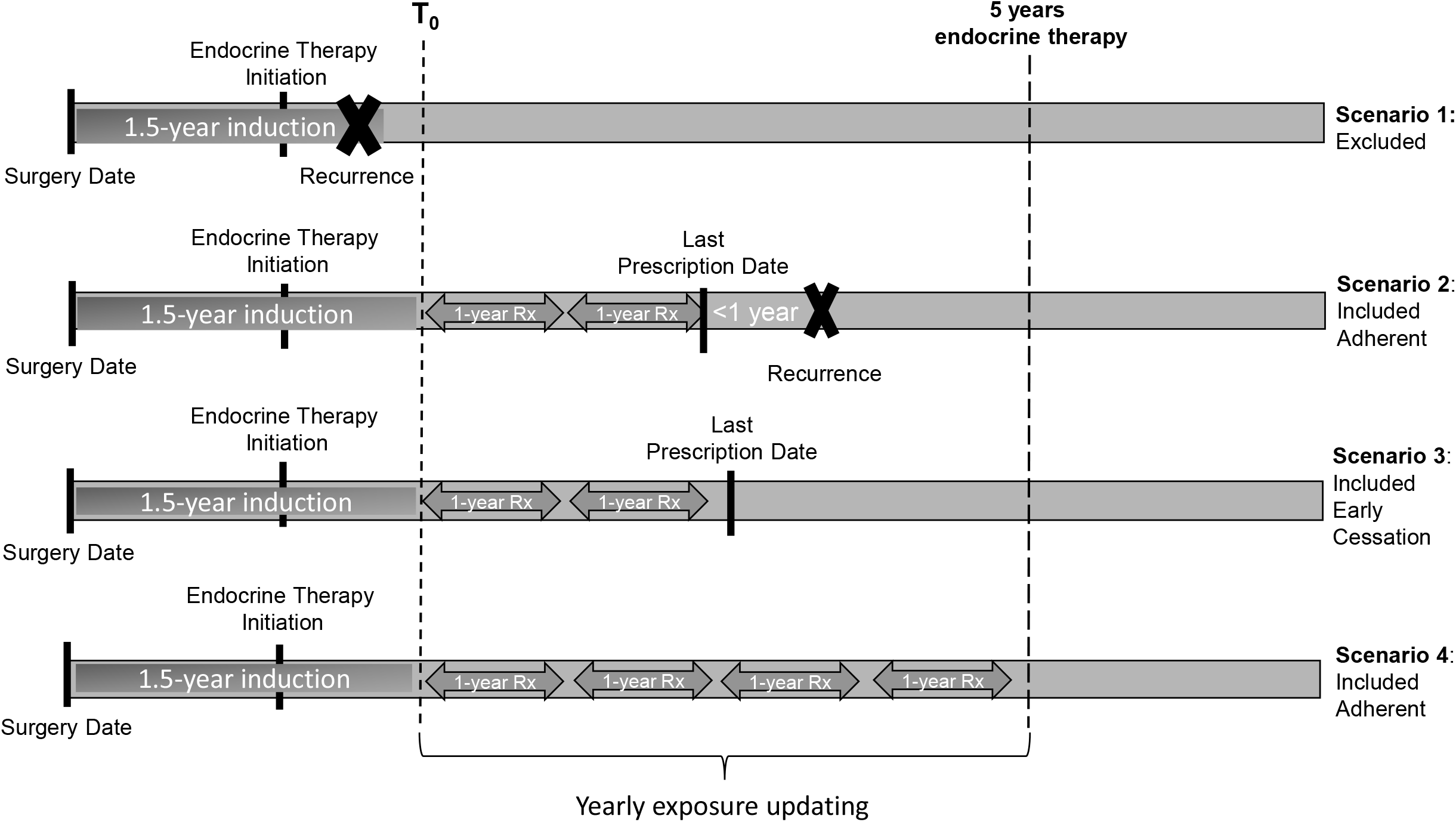
Illustration of cohort entry (surgery date), induction period (1.5 years), initiation of follow-up (T_0_), time-varying exposure status, and follow-up for study cohort.

### Covariates

Using the DBCG registry, the ProBe CaRe premenopausal cohort collected information about the first primary breast cancer diagnosis, including demographic, tumor, and treatment characteristics. This information included the date of surgery, type of surgery (mastectomy or breast conserving surgery), age at diagnosis, tumor size, tumor grade, lymph node status, human epidermal growth factor receptor-2 (HER2) receptor expression (available after 2007), stage—derived from tumor size (T), lymph node status (N), and metastases (M)—, radiation therapy, anti-HER2 therapy, and adjuvant chemotherapy. Among baseline patient characteristics, potential confounders include comorbidities (Charlson comorbidity index), socioeconomic status (median household income in tertiles), marital status (single, married), and education (high school, college, or graduate school). Medication persistence—defined as the continued per-protocol use—can be influenced by time-varying covariates. As such, incident comorbid diagnoses during follow-up, initiation of concomitant medication, and switching from tamoxifen to aromatase inhibitors were treated as potential time-varying confounders.

### Statistical Analysis

For descriptive analysis, we calculated the frequencies and proportions of patient demographic factors, tumor characteristics, treatments received, concomitant medications, and comorbidities within strata of tamoxifen early discontinuation status. We then evaluated the association between early discontinuation and recurrence using two complementary approaches.

First, we used pooled logistic regression models to estimate the time-dependent hazard ratios (HRs) and 95% confidence intervals (CI) for the association between early discontinuation of endocrine therapy and breast cancer recurrence.^25^ The pooled logistic regression models allow for a flexible baseline hazard and produce estimates that approximate the average rate ratio, similar to a Cox proportional hazards model with a time-dependent exposure.^26^ The exposure is time-varying to allow for treatment discontinuation at different timepoints over the five or more years of prescribed endocrine therapy. We updated the exposure annually after endocrine treatment initiation. We present multivariable-adjusted HRs and 95% CI accounting for potential baseline confounders.

Realizing that the experience of women who discontinue adjuvant endocrine therapy is not likely exchangeable with the experience of women who continue on therapy, for the second approach, we used G-computation to estimate the effect of early discontinuation of tamoxifen therapy on breast cancer recurrence adjusted for baseline tumor, treatment, and patient-level characteristics as well as time-varying covariates.^27^ To perform this analysis, we first calculated stabilized time-varying weights for each year of treatment. The weights for each patient were calculated from multivariable logistic regression models to estimate the probability of early discontinuation in that time interval based on baseline characteristics and time-varying factors, namely concomitant medication use and comorbidity diagnoses during follow-up. We used stabilized weights, computed by dividing the probability of discontinuing therapy given baseline characteristics by the probability of discontinuing therapy given baseline characteristics and time-varying covariates.^27^ These stabilized weights were then used as time-varying weights in a pooled logistic regression model to estimate the HR for the association between early treatment discontinuation and breast cancer recurrence. We calculated the accompanying 95% CI using nonparametric bootstrapping with 50 resamples.^28^

In a sensitivity analysis, we used a longer period to define early discontinuation. We allowed for 1.5 years between last date of prescription and end of follow-up, still considering a 6-month supply of endocrine therapy. This sensitivity analysis assumes that any event that occurred within one year of stopping endocrine therapy would not be due to early discontinuation of therapy.

## Results

Of the 4,503 premenopausal women included in the cohort, 1001 (22%) discontinued endocrine therapy over the course of follow-up (**Table 1**). The annual proportion of patients who discontinued therapy was lowest in the first year (2.6%), and gradually increased each year, culminating in a 6.7% discontinuation proportion in the 5^th^ year of therapy. Among those who discontinued endocrine therapy, 202 (20%) had a recurrence; among those who complete therapy, 388 (11%) had a recurrence (**Figure 2**). Compared with women who completed endocrine therapy, those who discontinued were less likely to have received chemotherapy (86% vs. 92%) or radiation therapy (49% vs. 55%). Women who discontinued were also more likely to have had a comorbid condition at diagnosis (11% vs 8.4%), have had a lower household income (42% vs. 31%), have been single during follow-up (36% vs. 32%), and have had a high school level of education (22% vs 19%).

**TABLE 1.** Distribution of clinical and tumor characteristics by early discontinuation versus completion of tamoxifen therapy among 4,503 premenopausal women with ER-positive breast cancer in a population-based cohort of premenopausal women diagnosed with first primary breast cancer, ProBe CaRe study (2002–2011).

| Patient and tumor characteristics |  | Early Discontinuation |  | Complete Therapy |  |
| --- | --- | --- | --- | --- | --- |
|  |  | N | % | N | % |
| **Total** |  | 1001 | (22) | 3502 | (78) |
| **Recurrence** |  |  |  |  |  |
|  | Yes | 202 | (20) | 388 | (11) |
|  | No | 799 | (80) | 3144 | (89) |
| **Switched from Tamoxifen to AI** |  |  |  |  |  |
|  | TAM to AI | 306 | (30) | 1502 | (42) |
|  | TAM only | 705 | (70) | 2087 | (58) |
| **Age at diagnosis** |  |  |  |  |  |
|  | <35 | 64 | (6.4) | 146 | (4.2) |
|  | 35–39 | 108 | (11) | 366 | (10) |
|  | 40–44 | 228 | (23) | 877 | (25) |
|  | 45–49 | 360 | (36) | 1276 | (36) |
|  | 50+ | 241 | (24) | 837 | (24) |
| **Stage at diagnosis** |  |  |  |  |  |
|  | Stage I | 277 | (28) | 885 | (25) |
|  | Stage II | 506 | (51) | 1935 | (55) |
|  | Stage III | 214 | (21) | 664 | (19) |
| **Tumor Diameter** |  |  |  |  |  |
|  | < 2cm | 514 | (51) | 1834 | (52) |
|  | 2 to <5cm | 436 | (44) | 1488 | (42) |
|  | ≥ 5cm | 49 | (4.9) | 180 | (5.1) |
| **Number of metastatic lymph nodes** |  |  |  |  |  |
|  | 0 | 392 | (39) | 1280 | (37) |
|  | 1 | 235 | (24) | 903 | (26) |
|  | 2 | 114 | (11) | 455 | (13) |
|  | 3+ | 258 | (26) | 854 | (24) |
| **Histologic grade** |  |  |  |  |  |
|  | I | 199 | (21) | 742 | (21) |
|  | II | 536 | (57) | 1815 | (57) |
|  | III | 204 | (22) | 708 | (22) |
|  | Unknown | 62 |  | 237 |  |
| **HER2 status** |  |  |  |  |  |
|  | HER2– | 591 | (81) | 2243 | (83) |
|  | HER2+ | 141 | (19) | 455 | (17) |
|  | Unknown/Not measured | 269 |  | 804 |  |
| **Type of primary surgery** |  |  |  |  |  |
|  | Mastectomy | 436 | (44) | 1549 | (44) |
|  | Lumpectomy | 565 | (56) | 1953 | (56) |
| **Chemotherapy** |  |  |  |  |  |
|  | Yes | 858 | (86) | 3219 | (92) |
|  | No | 143 | (14) | 283 | (8.1) |
| **Radiation therapy** |  |  |  |  |  |
|  | Yes | 491 | (49) | 1928 | (55) |
|  | No | 514 | (51) | 1574 | (45) |
| **Anti-HER2 therapy** |  |  |  |  |  |
|  | Yes | 82 | (8.2) | 383 | (11) |
|  | No | 919 | (92) | 3119 | (89) |
| **Charlson Comorbidity Score** |  |  |  |  |  |
|  | 1+ | 111 | (11) | 292 | (8.4) |
|  | 0 | 890 | (89) | 3210 | (92) |
| **Comorbidities during treatment** |  |  |  |  |  |
|  | 1+ | 95 | (9.5) | 349 | (10) |
|  | 0 | 906 | (91) | 3153 | (90) |
| **Statin Use** |  |  |  |  |  |
|  | Yes | 69 | (6.9) | 185 | (5.6) |
|  | No | 932 | (93) | 3317 | (95) |
| **Medication Initiation during treatment** |  |  |  |  |  |
|  | Yes | 610 | (61) | 2029 | (58) |
|  | No | 391 | (39) | 1473 | (42) |
| **Socioeconomic Factors** |  |  |  |  |  |
| **Household Income** |  |  |  |  |  |
|  | 3rd tertile | 281 | (28) | 1223 | (35) |
|  | 2nd tertile | 297 | (30) | 1206 | (34) |
|  | 1st tertile | 418 | (42) | 1068 | (31) |
| **Marital Status** |  |  |  |  |  |
|  | Married | 637 | (64) | 2406 | (69) |
|  | Single | 364 | (36) | 1096 | (31) |
| **Education** |  |  |  |  |  |
|  | Higher Education | 345 | (35) | 1408 | (41) |
|  | College | 426 | (43) | 1408 | (41) |
|  | High School | 219 | (22) | 659 | (19) |

Abbreviations: AI=aromatase inhibitors; HER2= human epidermal growth factor receptor 2; TAM=tamoxifen

**Figure 2:**
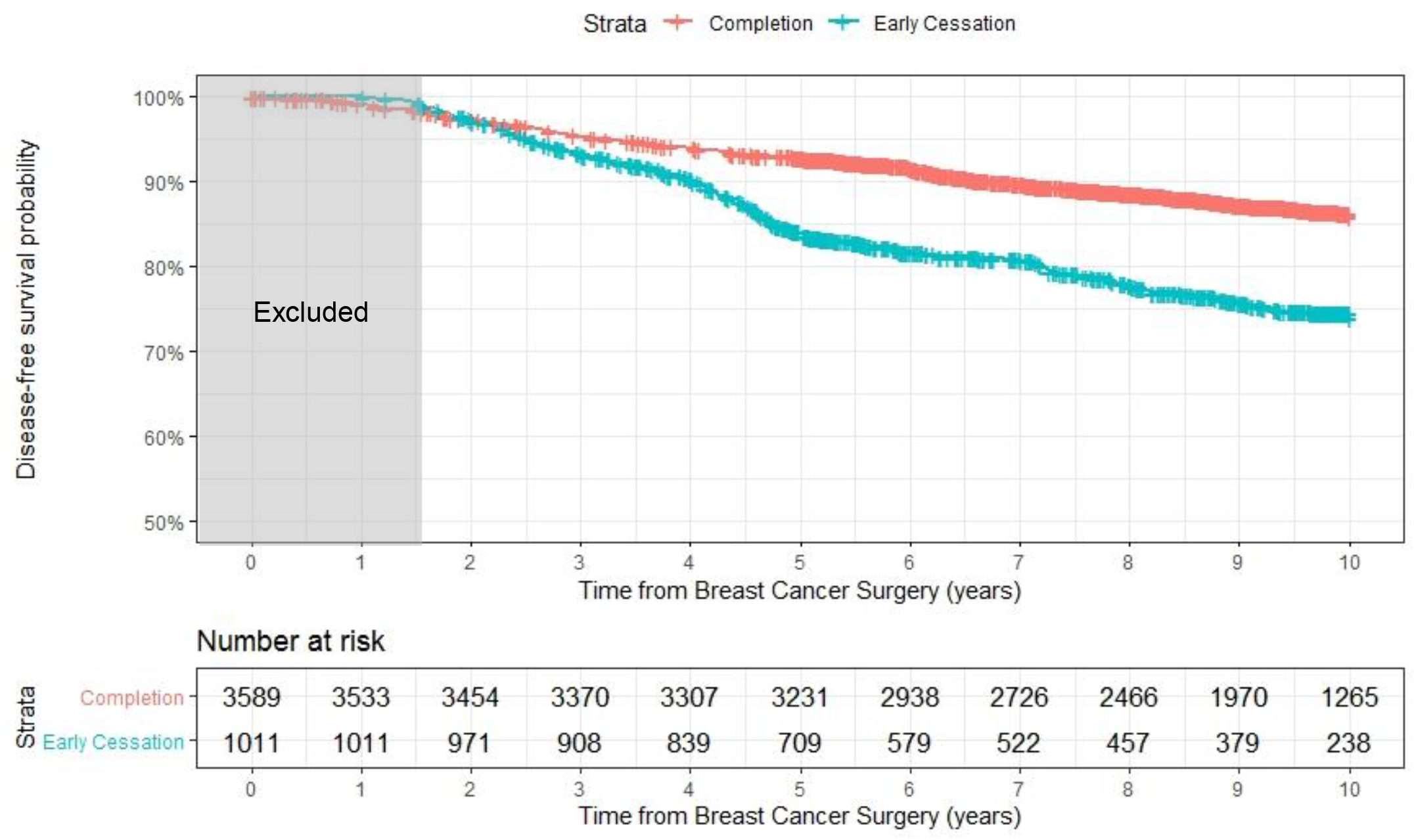
Kaplan-Meier survival functions by early discontinuation or completion of endocrine therapy among 4,600 premenopausal women diagnosed with breast cancer in Denmark (2002–2011) in the ProBe CaRe cohort.

In multivariable-adjusted models, early discontinuation of endocrine therapy was associated with an increase in the estimated rate of recurrence (HR=1.63, 95% CI: 1.30, 2.04) [**Table 2**]. Similarly, results from the G-computation approach (using time-varying weights) indicated that women who discontinued endocrine therapy had 1.67 times the estimated rate of recurrence compared with women who continued endocrine therapy (95% CI: 1.25, 2.14). The crude results were comparable to the above approaches, suggesting little confounding by measured baseline or time-varying confounders (HR=1.59, 95% CI: 1.27, 2.00).

**Table 2:** Multivariable analysis and sensitivity analysis for the association between early treatment discontinuation and breast cancer recurrence among 4,503 premenopausal women diagnosed with breast cancer in Denmark (2002–2011) in the ProBe CaRe cohort.

| Model | Multivariable Analysis |  | Sensitivity Analysis<sup>a</sup> |  |
| --- | --- | --- | --- | --- |
|  | HR | 95%CI | HR | 95%CI |
| Unadjusted | 1.59 | (1.27, 2.00) | 1.44 | 1.12, 1.83) |
| Adjusted<sup>b</sup> | 1.63 | (1.30, 2.04) | 1.49 | (1.15, 1.92) |
| G Computation<sup>c</sup> | 1.67 | (1.25, 2.14) | 1.41 | (1.15, 1.67) |

<sup>a</sup>Allows 1.5 years from last prescription and end of follow-up
<sup>b</sup>Baseline confounders include: age, stage, surgery type, radiation therapy, chemotherapy, statin use, comorbidity index, income, marital status, and education
<sup>c</sup>Time-varying confounders include: switch from tamoxifen to aromatase inhibitors, new comorbidity diagnosis, new medication use
Abbreviations: HR=hazard ratio; CI=confidence interval

The sensitivity analyses, which defined completion of adjuvant endocrine therapy as four years of therapy or continuation of therapy for up to one year before a recurrence, yielded similar, although attenuated estimates (**Table 2**).

## Discussion

This study is the first to quantify the effect of early discontinuation of adjuvant endocrine therapy on breast cancer recurrence in a population-based cohort of premenopausal women diagnosed with breast cancer. We report that 22% of premenopausal women discontinued endocrine therapy before completing five years of recommended treatment. The estimated rate of breast cancer recurrence in this study was about 1.7-fold higher among women who discontinued endocrine therapy compared with those who completed their course of treatment. Among women who did not complete adjuvant endocrine therapy, most (57%) discontinued taking medication in the last two years of treatment.

Trial results have reported that 5 years of tamoxifen therapy, compared with placebo, reduced the risk of recurrence by nearly one-half among premenopausal women diagnosed with ER-positive breast cancer.^29^ This result suggests that lack of tamoxifen therapy among premenopausal women diagnosed with ER-positive breast cancer would nearly double the risk of recurrence compared with tamoxifen therapy of appropriate duration. A recent systematic review reported that 15%–20% of women discontinued tamoxifen therapy in the first year of therapy and 31%–60% discontinued by the end of 5 years.^30–34^ Although the review primarily included studies of U.S. populations—in which a greater proportion of women than in Denmark discontinue endocrine therapy—the pattern of discontinuation over the first 5 years of therapy is comparable to the pattern observed in our study. A meta-analysis of trials comparing the benefit of 5 years vs 1–2 years of tamoxifen therapy reported 0.82 times the rate of recurrence.^29^ Therefore the expected increase in the rate of recurrence comparing 2 vs 5 years of tamoxifen therapy would be approximately 1.22 (1/0.82).^29^ The estimated effect from our study is further from the null than this expectation, likely due to a combination of factors. First, there is a difference in target populations, as the trials were conducted among both pre- and postmenopausal breast cancer patients, whereas our study is restricted to premenopausal women. Second, most women in our study who completed 5 years of tamoxifen therapy continued on therapy, given the change in guidelines from 5 to 10 years of therapy in 2013.^35^ Third, our study estimated the rate ratio, which will be further from the null than the risk ratio reported from trials. Finally, we excluded recurrences in the first 1.5 years after diagnosis, which were included in the trial results. This exclusion reduces the number of events, and ratios of rarer events are often larger than ratios of more common events.^36^ Even though these women would have received some benefit from initiating therapy, evidence clearly supports the need for completion of the 5 years of tamoxifen therapy—and even extending to 10 years—to acquire the full preventive benefit.

Before the end of 5 years of adjuvant endocrine therapy, 22% of women in our cohort discontinued treatment. This estimate is substantially lower than those from previous studies conducted in the U.S.;^19^ however, the proportion is slightly larger than the corresponding 16% of postmenopausal women who discontinued treatment reported in an earlier Danish study.^20^ This difference may be expected because some previous studies have shown that younger women are less likely to complete adjuvant endocrine therapy than older women.^10^ Similar to U.S.-based studies, we observed that women who discontinued therapy were more likely to report a lower household income, be unmarried, and have high school rather than third-level education, compared with women who completed therapy.^37^ Income, education, and healthcare access gaps are more pronounced in the U.S. than in Denmark, which is also correlated with healthcare access, and may partially explain the differences observed between our study and the review.^38,39^ The BIG 1-98 study—a double-blind trial that randomized postmenopausal women to tamoxifen or aromatase inhibitors—reported tamoxifen discontinuation rates that were consistent with those observed in the current study (19%).^40^ In BIG 1-98, among postmenopausal women who were randomized to the tamoxifen arm, adverse events such as thromboembolic complications were the most common causes for treatment discontinuation.^40^ In the clinical setting, women who are at risk for or who experience thromboembolic complications are eligible to switch to aromatase inhibitors, which carry about half the risk of thrombotic events compared with tamoxifen.^41,42^ Still, adverse side effects are often reported as a reason that patients stop taking tamoxifen.^12,43^

Our results are comparable to those observed in other studies that were not restricted to premenopausal women. A study using a Kaiser Permanente patient database reported that early discontinuation was associated with higher all-cause mortality (HR 1.26, 95% CI: 1.09, 1.46).^44^ In a Danish population-based cohort of postmenopausal women diagnosed with breast cancer between 1996 and 2004, those who discontinued therapy had an increased hazard of recurrence (HR=1.45, 95% CI: 1.14, 1.85).^20^ The associations observed in our study are more pronounced, underscoring the importance of endocrine therapy completion among premenopausal women to prevent or delay a recurrence.

There are several limitations to consider. First, as with all population-based studies of medication use, we had information on prescriptions dispensed at each follow-up visit but could not be certain that endocrine therapy was taken as prescribed. It is likely that women who discontinued therapy before the recommended 5 years of treatment were also less likely to take the medication as indicated, even when they filled a prescription. Second, since 2013, the Danish guidelines recommend that tamoxifen be prescribed to premenopausal women for up to 10 years.^7,35^ We observed that, among women who completed at least 5 years of adjuvant endocrine therapy, 11% (n=405) completed 10 years of treatment, and 71% (n=2,565) continued taking medication for more than 5 years. The proportion of women receiving more than 5 years of adjuvant endocrine therapy increased as a function of year of diagnosis between 2002 and 2011. Given that our study population included women diagnosed with breast cancer through 2011, and with follow-up available until August 31, 2019, we were unable to examine the effect of discontinuation of tamoxifen therapy at 5 years compared with continuation for up to 10 years. Third, we were unable to capture menopausal transition. We did observe that 40% of those who completed therapy and 30% of those who did not complete therapy switched from tamoxifen to an aromatase inhibitor. This suggests that a large proportion of women transitioned to menopause, but women may also switch type of therapy due to side effects. Finally, women who discontinued therapy were more likely to have had a recurrence within the first 5 years following their diagnosis compared with those who completed therapy (70% vs. 52%). This may reflect an innate resistance to adjuvant endocrine therapy.^45–47^ We excluded women with less than 1.5 years of follow-up or a recurrent event within the first 1.5 years following diagnosis to help mitigate this potential bias. Still, the hazard of recurrence is highest in the first 2–3 years after diagnosis, and the prevalence of discontinuation increases over time.^48^ The combination of eliminating the first 1.5 years of follow-up and a discontinuation proportion that increases over time suggests that the association observed in this study pertains to recurrences that occur after the initial high hazard period. If we assume that recurrences in the high hazard period reflect an innate resistance of the cancer rather than persistence of endocrine therapy, then our results are unbiased. However, future analyses might consider analyzing the results stratified based on year of discontinuation to look specifically at the influence of endocrine therapy on this early period of high recurrence hazard.

In conclusions, our results highlight the importance of clinical follow-up and behavioral interventions that support persistence of adjuvant endocrine therapy to prevent a recurrent event.

## Data Availability

Data are available within the paper. Individual level datasets are not publicly available in accordance with the Danish Act on Processing of Personal Data (https://www.datatilsynet.dk/english/the-act-on-processing-of-personal-data/read-the-act-on-processing-of-personal-data/compiled-version-of-the-act-on-processing-of-personal-data/). Interested researchers can contact the corresponding author for data requests.

